# Sensitivity Analysis on Predictive Capability of SIRD Model for Coronavirus Disease (COVID-19)

**DOI:** 10.1101/2020.11.21.20236083

**Authors:** Ahmad Sedaghat, Fadi Alkhatib, Seyed Amir Abbas Oloomi, Mahdi Ashtian Malayer, Amir Mosavi

## Abstract

SIR model is one of the simplest methods used in prediction of endemic/pandemic outbreaks. We examined SIRD model for development of COVID-19 in Kuwait which was started on 24 February 2020 by 5 patients in Kuwait. This paper investigates sensitivity of SIRD model for development of COVID-19 in Kuwait based on duration of progressed days of data. For Kuwait, we have fitted SIRD model to COVID-19 data for 20, 40, 60, 80, 100, and 116 days of data and assessed sensitivity of the model with number of days of data. The parameters of SIRD model are obtained using an optimization algorithm (lsqcurvefit) in MATLAB. The total population of 50,000 is equally applied for all Kuwait time intervals. Results of SIRD model indicates that after 40 days the peak infectious day can be adequately predicted; althogh, error percentage from sensetivity analysis indicates that different exposed population sizes are not correctly predicted. SIRD type models are too simple to robustly capture all features of COVID-19 and more precise methods are needed to tackle nonlinear dynamics of a pandemic.

## Introduction

Since the outbreak of COVID-19 in Wuhan, China in December 2019, 215 countries worldwide reported the pandemic cases of COVID-19 summing total 9,655,329 diagnosed cases, total 488,136 death cases, and total 5,244,462 recovered cases on 26 June 2020 [1].

Kuwait reported total 42,788 diagnosed, current 9,082 under treatment, 152 critical, 339 deceased, 33,367 recovered, and 23 quarantined on 26 June 2020 by the ministry of health (MOH) [2]. Kuwait government have already removed full lockdown except for some highly susceptible areas and taken steps to ease on most of closures across the country.

Accurate prediction of COVID-19 development is proved to be very difficult in many countries due to unstable nonlinear dynamics of the pandemic in these countries. To show uncertainty of our prediction on dynamics of COVID-19 reported in [3], we have assessed our optimized SIRD model based on time intervals of COVID-19 developments.

In this paper, we have used SIRD model optimized by an optimization algorithm (lsqcurvefit) in MATLAB for fitting the model with COVID-19 data for several consequence time intervals in Kuwait and UAE. We have calculated the goodness of fit using the coefficient of determination (R^2^). Sensitivity of results to choose of number of COVID-19 data are presented and discussed for the pandemic outbreak in Kuwait and UAE and finally conclusions of the work are drawn.

## Methodology

### SIRD Model

SIR model is the simplest model used in epidemic/pandemic studies which is expressed by a set of consequence ordinary differential equations (ODE) introduced by Kermack andMcKendrick [4]. Details of 3 set of ODEs can be found in numerous publications based on susceptible (S) cases, active infected cases (I), and removed cases (R) including recovered and death [5, 6]. We used SIRD model which includes one more equation on number of death (D) as follows [7]:

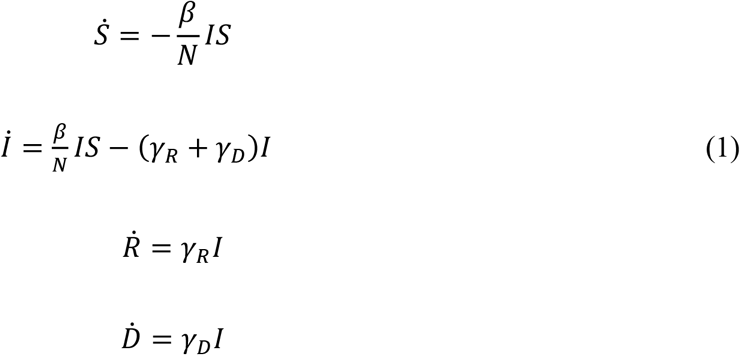

In equation (1), *S* is the susceptible, *I* is the active infected, *R* is the recovered, and *D* is the death populations. Time derivatise are shown by over-dot; e.g.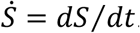. The transmission rate *β*, the recovery rate *γ*_*R*_, and the death rate *γ*_*D*_ are unknown SIR model parameters that are obtained by fitting COVID-19 data using the curve-fitting optimization algorithm (lsqcurvefit) in MATLAB. The removing rate is obtained by *γ* = *γ*_*R*_ + *γ*_*D*_. Re-production number *R*_0_ = *β*/*γ* is an important characteristic parameter on dynamics of SIR model. Some studies suggest that if *R*_0_ < 1 then the nonlinear dynamics of SIR model is stable and predictions are close to reality [8]. But, *R*_0_ > 1 then the dynamical system of SIR ODE equations is unstable. In SIR model, total population *N* is considered as constant.

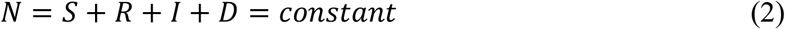

We should differentiate between total active infected (*I*) and total infected cases (*IC*) as follows:

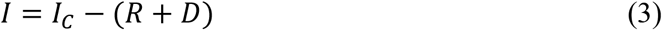

An example of initial conditions to solve SIR model in Kuwait is as follows:

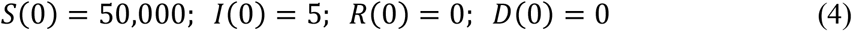

Optimized SIR model parameters are obtained for all time intervals of COVID-19 data in Kuwait using initial conditions (4) in MATLAB using the algorithm (fminsearch).

### Goodness of fit technique

#### Regression coefficient

Regression coefficient (R^2^) is applied for each of four populations, susceptible, infected, recovered, and death to check goodness of fitting SIR model with COVID-19 data. The regression coefficient is a good measure on dealing with futurism prediction. The regression coefficient (R^2^) compares predicted values (*y*) against actual data (*x*) as follows [9]:

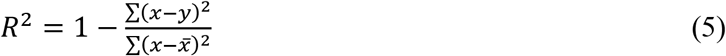

In equation (6), 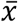 is the average of COVID-19 data values. The regression coefficient (R^2^) close to unity shows best fit.

### Sensitivity analysis

Having actual peak day values, SIRD model predictions using different time duration set data can be assessed to determine sensitivity of the model to size of COVID-19 data. A comparison of actual peak day values (*x*) and predicted values (*y*) are obtained in percentage of error as follows [10]:

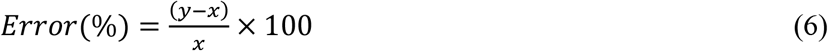

Sensitivity of SIRD model is obtained using equation (6) on time duration of COVID-19 data values. Closer error to zero percentage means less sensitivity of SIRD model to data size.

### SIRD Predictive Results for Kuwait

Results of SIRD model using COVID-19 data for 20, 40, 60, 80, 100, and 116 days with optimized parameters from 24 February to 19 June 2020 are shown in Fig. 1. Table 1 summarizes optimized parameters obtained corresponding to cases in Fig. 1. It is observed that 20 days data cannot provide any coefficient of regressions for different population due to lack of data. For instance, there was zero death cases during the first 20 days period so it is not possible to determine the regression coefficient. For 40- and 60-days interval, the death data was still insufficient to provide a meaningful accuracy.

**Table 1:** Trends of optimized SIRD model parameters and accuracy of results for COVID-19 in Kuwait (19 June 2020).

| No. of days | Growth rate ($\beta$) | Recovery rate ($\gamma_R$) | Death rate ($\gamma_D$) | Removing rate ($\gamma$) | Reproduction number ($R_0$) | $R^2(S)$ | $R^2(I)$ | $R^2(R)$ | $R^2(D)$ |
| --- | --- | --- | --- | --- | --- | --- | --- | --- | --- |
| 20 | 0.18311 | 0.01363 | 0.00435 | 0.01798 | 10.18 | - | - | - | - |
| 40 | 0.14507 | 0.03524 | 0.00215 | 0.03739 | 3.88 | 0.81409 | 0.63761 | 0.95812 | - |
| 60 | 0.12943 | 0.02308 | 0.00156 | 0.02464 | 5.25 | 0.96106 | 0.93959 | 0.98122 | - |
| 80 | 0.13963 | 0.04158 | 0.00137 | 0.04295 | 3.25 | 0.99150 | 0.97154 | 0.98236 | 0.57438 |
| 100 | 0.13399 | 0.03576 | 0.00154 | 0.0373 | 3.59 | 0.99701 | 0.98373 | 0.98172 | 0.70984 |
| 116 | 0.15188 | 0.05501 | 0.00099 | 0.056 | 2.71 | 0.99870 | 0.91169 | 0.96943 | 0.78406 |

**Figure 1:**
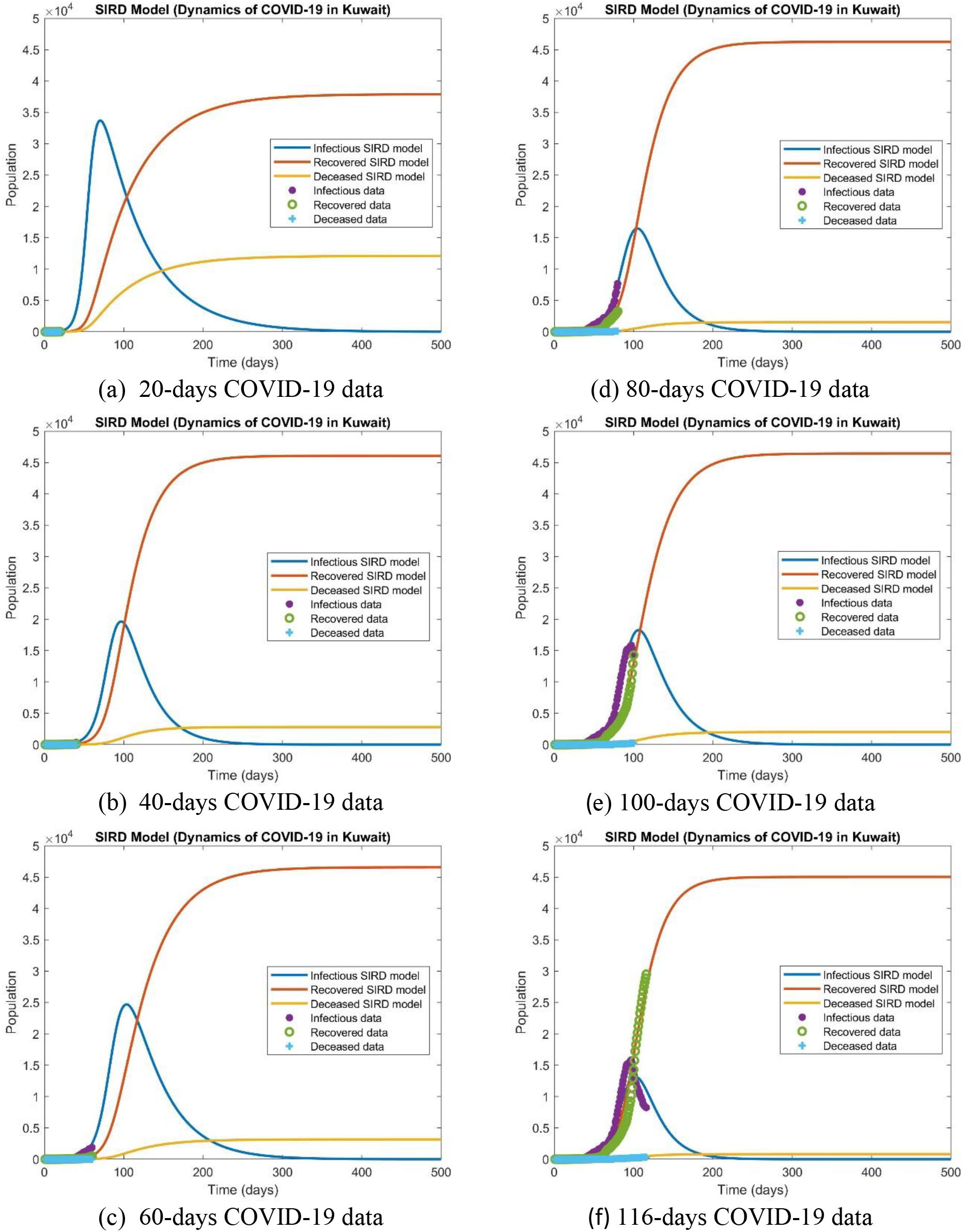
Optimized SIRD model predictions for dynamics of COVID-19 in Kuwait (19 June 2020).

Figure 2 compares values of susceptible, active infected, recovered, deceased, and total infected using various time duration of COVID-19 data for peak day of infection. As shown in Fig. 2, with 20, 40, 60, 80, 100, and 116 days of data the peak infection dates 5/5, 1/6, 7/6, 9/6, 10/6, and 6/6/2020 were predicted, respectively. A linear trend is observed in all of these predictions for different populations; yet, we do not get precise solution.

**Figure 2:**
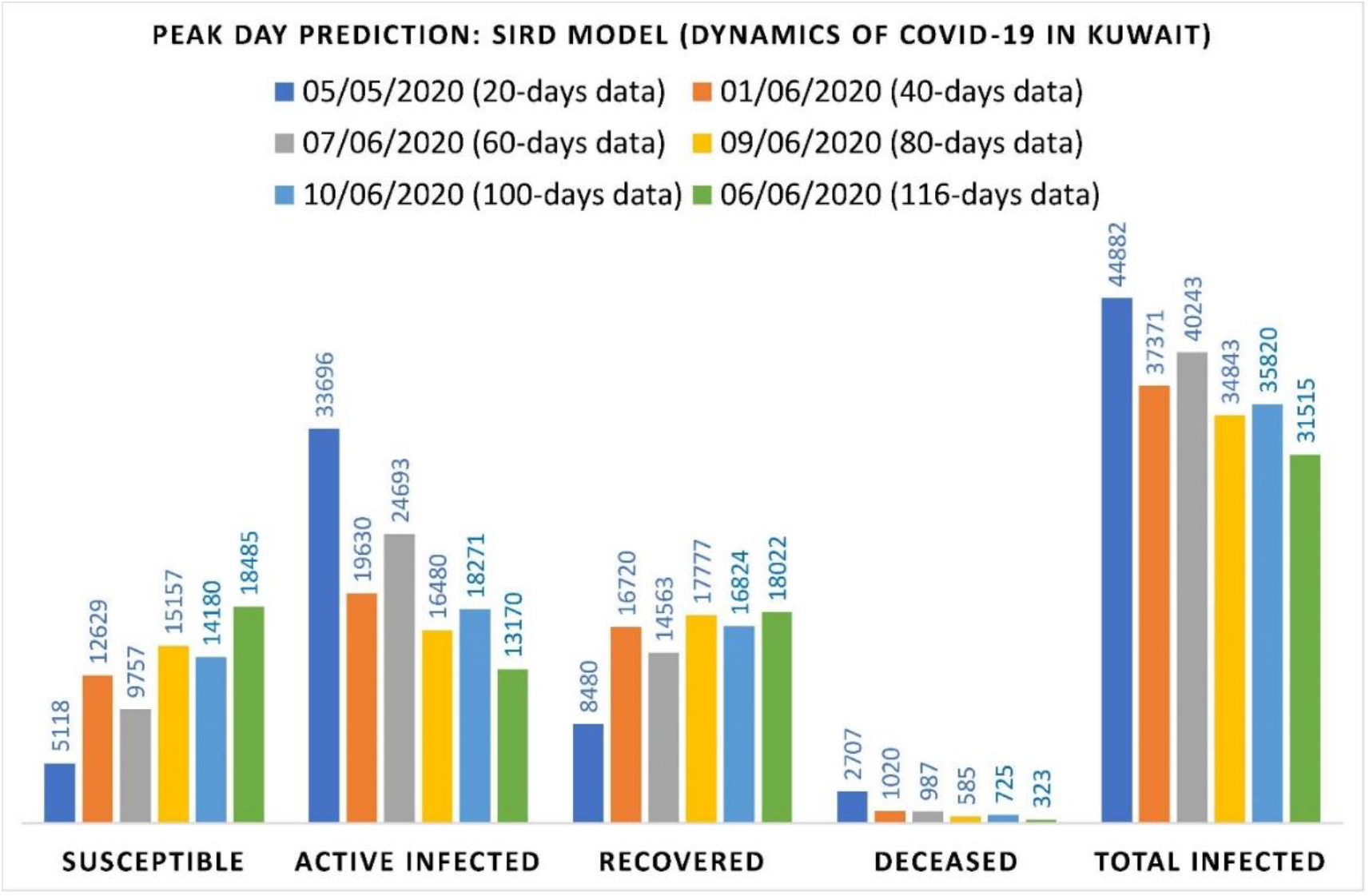
Peak infection day predictions using optimized SIRD model using 20, 40, 60, 80, 100, and 116 COVID-19 data-sets (19 June 2020)

Figure 3 shows the trends of these solutions as COVID-19 days progressed. No convergence to certain limits is observed in this figure. Actual peak day of infection was observed on 30 May 2020 (96 days after 24 February 2020) according to worldometer [1].

**Figure 3:**
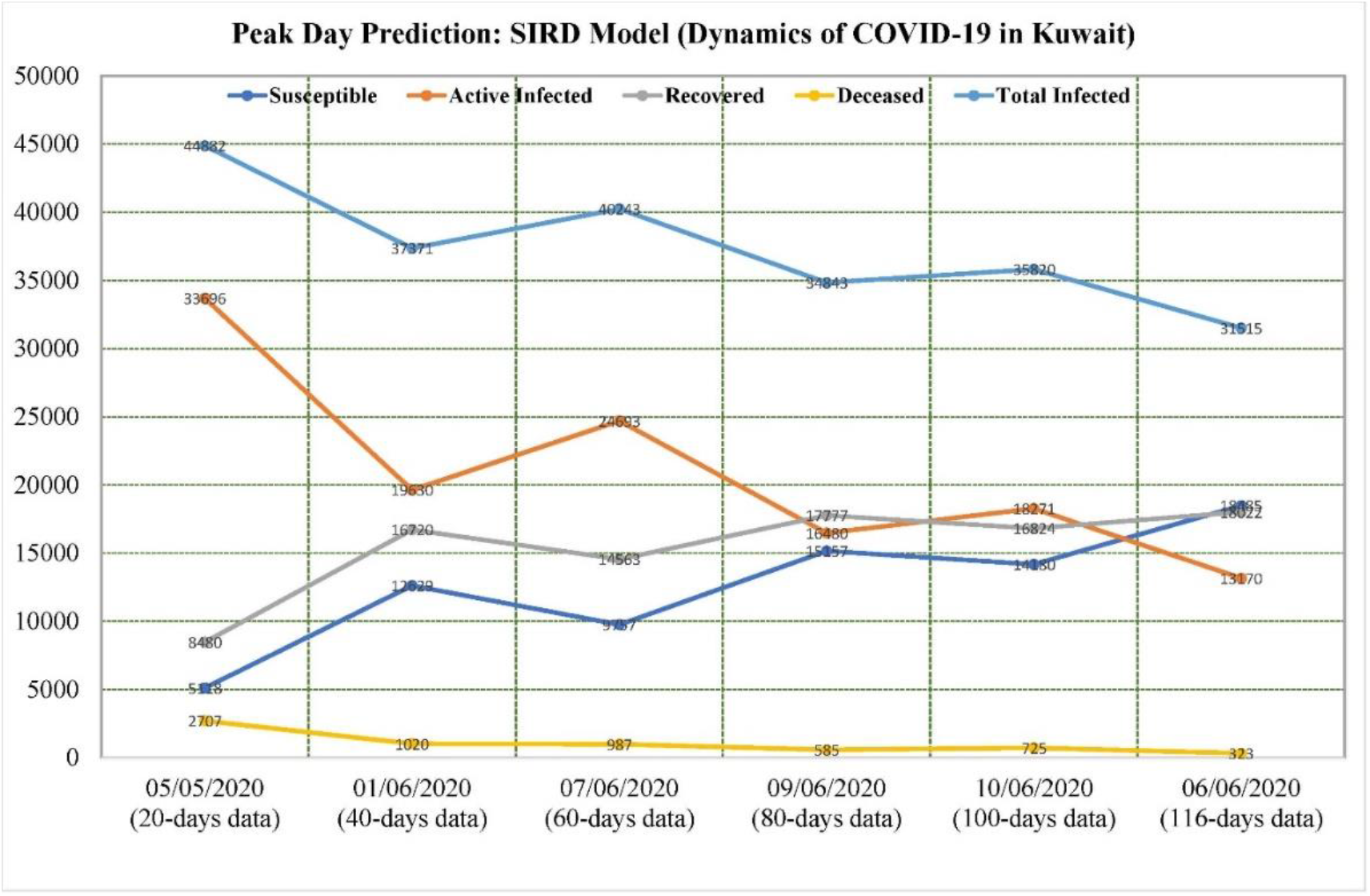
Population size trends on peak infection day using optimized SIRD model for 20, 40, 60, 80, 100, and 116 COVID-19 data-sets (19 June 2020).

The predicted and actual values of peak infectious day and corresponding errors using equation (6) are shown in Table 2. The lowest error is 17% on total infected cases using 116 days of COVID-19 data. The results of Table 2 show poor predictive capability of SIRD model.

**Table 2:** Sensitivity of optimized SIRD model results on accuracy of predicted peak infectious day of COVID-19 in Kuwait (19 June 2020).

| No. of days | Susceptible population (S) | Active Infected population (I) | Recovered population (R) | Deceased Population (D) | Total infected cases (I<sub>c</sub>) | Error (%) S | Error (%) I | Error (%) R | Error (%) D | Error (%) I<sub>c</sub> |
| --- | --- | --- | --- | --- | --- | --- | --- | --- | --- | --- |
| 20 | 5118 | 33696 | 8480 | 2707 | 44882 | -79 | 113 | -17 | 1220 | 71 |
| 40 | 12629 | 19630 | 16720 | 1020 | 37371 | -47 | 24 | 65 | 397 | 43 |
| 60 | 9757 | 24693 | 14563 | 987 | 40243 | -59 | 56 | 43 | 382 | 54 |
| 80 | 15157 | 16480 | 17777 | 585 | 34843 | -36 | 4 | 75 | 185 | 33 |
| 100 | 14180 | 18271 | 16824 | 725 | 35820 | -40 | 15 | 66 | 254 | 37 |
| 116 | 18485 | 13170 | 18022 | 323 | 31515 | -22 | -17 | 77 | 57 | 20 |
| *96 | 23808 | 15831 | 10156 | 205 | 26192 | 0 | 0 | 0 | 0 | 0 |

\* Actual data on peak day

## Conclusions

It is highly important we estimate accurately COVID-19 dynamics as early as possible. Here, we investigated sensitivity of SIRD model on predicting trends of COVID-19 in Kuwait. We optimized SIRD model parameters so that we get high resolution and best fit to COVID-19 data for any time duration of data. We used 20, 40, 60, 80, 100, and 116 days data separately to fit SIRD model with each time interval COVID-19 data. The following conclusion can be drawn:

- SIRD model have accurately predicted the peak day of infectious after 40 days development of COVID-19 within 1-10 June 2020 in Kuwait. The actual peak day was 30 May 2020.
- Sensitivity analysis of SIRD model on peak infectious day indicated that the model is not very accurate with error bands of 22-79% for susceptible, 4-113% for active infected, 17-77% for recovered, 57-1220% for deceased, and 20-71% for total infected.
- SIRD model can be considered as a rough model for predicting number of exposed populations and better models with more controlling factors may be needed.
- Trends of reducing the re-production number (R0) since outbreak of COVID-19 in Kuwait is promising indicator in slowing down the disease; but, R0>1 shows unstable dynamics of COVID-19 in Kuwait.
- High value of the re-production number (R0) is a bad indicator and may lead to second wave of COVID-19 infection in Kuwait. Results of active infected cases on 26 June 2020 might indicates increasing trends towards a second peak of infectious.

SIRD model is a useful tool for rough estimation of peak day of infectious for COVID-19 pandemic; yet, it is not precise method. Advanced versions of SIR model or other new models are needed with more controlling factors to find more precise predictions.

## Data Availability

data is online:
Worldometer, https://www.worldometers.info/world-population/kuwait-population/, accessed 26 June 2020.

